# The effect of psychosocial and healthcare interventions on hospitalisation in people with dementia: an umbrella review

**DOI:** 10.1101/2025.10.13.25337873

**Authors:** Connie Howard, Alba Fernández-Sanlés, Nawal Abukar, Daniel Davis, Claire Goodman, Sanjiv Gupta, Melanie Handley, Naaheed Mukadam, Carol Riddington, Kate Walters, Gill Livingston, Andrew Sommerlad

## Abstract

**Objective:** To evaluate the efficacy of psychosocial and healthcare interventions on reducing hospitalisation in people with dementia (PwD).

**Design:** Umbrella review of existing systematic reviews and meta-analyses.

**Search strategy:** MEDLINE, Embase, Cochrane, and CINAHL were searched until September 2025 for relevant keywords and medical subject headings.

**Inclusion criteria:** Peer-reviewed systematic reviews with or without meta-analyses examining the effect of any psychosocial or healthcare intervention compared to a suitable control group on any measure of hospitalisation in people with dementia.

**Methods:** Two reviewers assessed eligibility, and eligible papers were grouped into categories and appraised by two reviewers using AMSTAR (A MeaSurement Tool to Assess systematic Reviews) version 2. We assessed certainty of evidence for each intervention type using GRADE (Grading of Recommendations Assessment, Development, and Evaluation).

**Results:** The search identified 25 systematic reviews, comprising 77 unique primary studies (47 randomised control trials, 30 non-randomised studies of interventions), totalling 1,483,077 participants. There was high-certainty evidence that case management and exercise programmes had no effect on hospitalisation of PwD. There was low certainty evidence that advance care planning (ACP) reduced hospital admissions. Moderate certainty evidence from one study suggested that including clinical pharmacists in multidisciplinary teams (MDTs) reduced medication-related hospital admissions.

**Conclusions:** The current evidence for psychosocial and healthcare interventions in reducing hospitalisation in dementia is insufficient to make strong recommendations. ACPs show promise in reducing hospitalisations, and clinical pharmacists in MDTs may reduce medication-related readmissions. As evidence-based recommendations are needed to reduce the burden of dementia-related hospitalisations, more robust research is needed, especially high-quality randomised control trials with greater detail and standardised checklists for interventions and outcomes to reduce heterogeneity and establish more confident clinical recommendations.

**Registration:** PROSPERO 2024 CRD42024604296

## Introduction

The number of people living with dementia (PwD) globally is expected to increase from 57 million in 2019 to 152 million by 2050 (1). As PwD have 42% increased risk of admission to acute hospital care compared with those without dementia, even after adjusting for physical comorbidities, there will be substantially increased hospital use (2). In the UK, PwD occupy one in six hospital beds, with £2.8 billion annual costs due to hospital inpatient admissions, and it is expected that by 2040 one quarter of hospital inpatients will have dementia (3). Once admitted, PwD stay in acute hospitals up to seven times longer than age-matched patients (4) and are at a 7-35% increased risk of readmission (5, 6). Admissions to acute hospitals can also have a significant negative impact on the health of PwD, including increased mortality (7–10) and frailty (9, 11), more inpatient complications (12, 13), and further cognitive and functional decline (8, 9, 13–16) with only 42% of PwD recovering to their pre-hospitalisation level of function (17). Acute hospitals are fast-paced environments that are usually not specifically designed to care for PwD (9, 18, 19), with staff who often report feeling inadequately prepared and supported to look after PwD (3, 20).

While some hospital admissions are essential and appropriate, 20-49% of hospitalisations of PwD are potentially preventable (4, 21, 22), so it is crucial to consider how to reduce these admissions. However, current evidence examining the effectiveness of psychosocial and healthcare interventions on reducing hospitalisation in PwD is spread across different intervention types, with unclear efficacy (23). Therefore, we conducted an umbrella review (24) to synthesise the evidence on the efficacy and quality of psychosocial and healthcare interventions aimed at reducing hospitalisation rates in PwD.

## Methods

We registered our umbrella review in PROSPERO (CRD42024604296) and report our umbrella review according to Preferred Reporting Items for Overviews of Reviews (PRIOR) guidelines (25) (See Appendix Table 3). We systematically searched the literature for published systematic reviews and meta-analyses that examined the association between psychosocial and healthcare interventions and hospitalisation (including hospital admission and readmission, length of stay, emergency department (ED) visits, and delayed discharge) in PwD.

### Information sources/ search strategy

We searched systematically across four electronic databases from inception —MEDLINE, Embase, Cochrane, and CINAHL — in November 2024, with no language or date restrictions. Searches were re-run on 15^th^ September 2025 to ensure recent reviews were included. Keywords and Medical Subject Headings terms such as “Hospitalization”, “Emergency Room visits”, and “Dementia” were combined with Boolean operators to identify relevant systematic reviews and meta-analyses. See Appendix for complete search strategy.

### Selection process

Duplicates were removed using Covidence software. Two reviewers (CH and NA) independently screened titles and abstracts, followed by full-text articles according to the predefined eligibility criteria. At each stage, disagreements were discussed with GL and AS to reach a consensus. Inter-rater reliability was assessed using Cohen’s Kappa, with values >0.81 considered almost perfect agreement, 0.61-0.80 substantial, 0.41-0.60 fair, and <0.40 none/slight agreement (26).

### Eligibility criteria

Eligibility criteria were established using the Population, Intervention, Comparator, Outcome, Study type (PICOS) framework. Systematic reviews were eligible for inclusion if they included: 1) PwD (population), 2) any psychological/ social support/ healthcare delivery interventions (intervention), 3) a suitable control group (control) and, 4) a measure of hospitalisation (outcome), 5) and was a systematic review (study type), defined as studies seeking to “collate all empirical evidence that fits pre-specific eligibility criteria to answer a specific research question” using explicit, predefined methods to minimise bias (27). Eligible study designs included randomised controlled trials (RCTs), non-randomised studies of interventions (NRSIs), observational studies with intervention analysis, quasi-experimental studies, and mixed-method studies.

Systematic reviews were excluded if they focused on mild cognitive impairment (MCI) or delirium, or if they analysed the effects of pharmacological interventions on hospitalisation. Studies were not eligible if they did not examine an intervention compared to a control. Therefore, qualitative studies, cross-sectional studies, editorials, letters, and abstracts were also excluded (24).

### Data extraction process

We created a custom document to extract the following data: author, publication year, location, methods (total relevant studies, number of RCTs and observational studies), population (total relevant sample size, inclusion and exclusion criteria, mean age and dementia severity information), intervention (intervention type, control group, setting, target and length), results for each outcome, and quality assessments. One reviewer (CH) independently extracted data from each review according to our custom template, and a second reviewer (NA) checked for any discrepancies. To visualise overlap in primary studies between reviews, two reviewers (CH and NA) created a citation matrix, which was coordinated according to the interventions used in each primary study.

Where a primary study intervention was categorised differently across reviews, one reviewer (CH) examined the primary study to determine the accurate intervention label and adjusted the matrix, accordingly, discussing uncertainties within the research team. Similarly, where individual study sample sizes differed across reviews, one reviewer (CH) assessed the primary study to determine its true sample size. Corresponding authors of systematic reviews were contacted (28, 29) for clarification if they used unpublished data which did not correspond with the primary study. See Appendix for citation matrix.

### Risk of bias assessment

Risk of bias was assessed using the AMSTAR II tool (A Measurement Tool to Assess Systematic Reviews) (30), which is designed to assess RCTs and NRSIs. Critical domains in the assessment include the pre-registration of review methods, comprehensive search strategies, justification of excluded studies, a satisfactory analysis of the risk of bias and publication bias, and the use of appropriate statistical combination methods. One reviewer (CH) independently rated all reviews according to these criteria and consulted AS and GL when unsure. See Appendix for full AMSTAR II ratings.

### Synthesis methods

We present our findings in a narrative synthesis. Due to study heterogeneity and a subsequent lack of meta-analyses in many systematic reviews, we could not conduct meta-analyses.

### Certainty assessment

The GRADE tool (Grading of Recommendations, Assessment, Development and Evaluation)(31) was used to assess the certainty of evidence. One reviewer (CH) assessed all included systematic reviews utilising this framework, with a second reviewer (AF-S) independently assessing 10% at random to check for consistency. According to this approach, all observational studies began as low-quality evidence, and RCTs began as high-quality evidence (32). Five criteria concern the downgrading of evidence: risk of bias, inconsistency, indirectness, imprecision, and publication bias, and three criteria gave the opportunity to upgrade the evidence: magnitude of effect, dose-response gradient, and plausible confounders. As some reviews included multiple interventions, with different effect estimates per intervention, the GRADE tool was applied to each intervention in each review so the ratings could make recommendations by intervention type (33). According to these criteria, each intervention within each review was then categorised as having either ‘high’, ‘moderate’, ‘low’, or ‘critically low’ certainty of evidence. Lower quality evidence did not detract from high quality findings.

### Public and patient involvement (PPI)

PPI members with experience caring for family members with dementia were involved in the umbrella review from its inception. Quarterly meetings were held with five members to discuss the review and its progress. PPI members assisted in creating a comprehensive search strategy, introducing lived experience to the interpretation of findings, and identifying crucial areas for future research. Two members (CR, SG) were involved as co-authors of the article

## Results

The initial search yielded 1,828 studies, with 136 additional articles found in September 2025 (see Figure 1 for flow of studies). After de-duplication, 1454 studies were eligible for screening. Following title and abstract screenings (n=1,454) and full text review (n=53), 25 reviews were selected for data extraction. Inter-rater agreement was substantial at the title and abstract screening stage (κ=0.72) and at full-text screening (κ =0.80).

**Figure 1:**
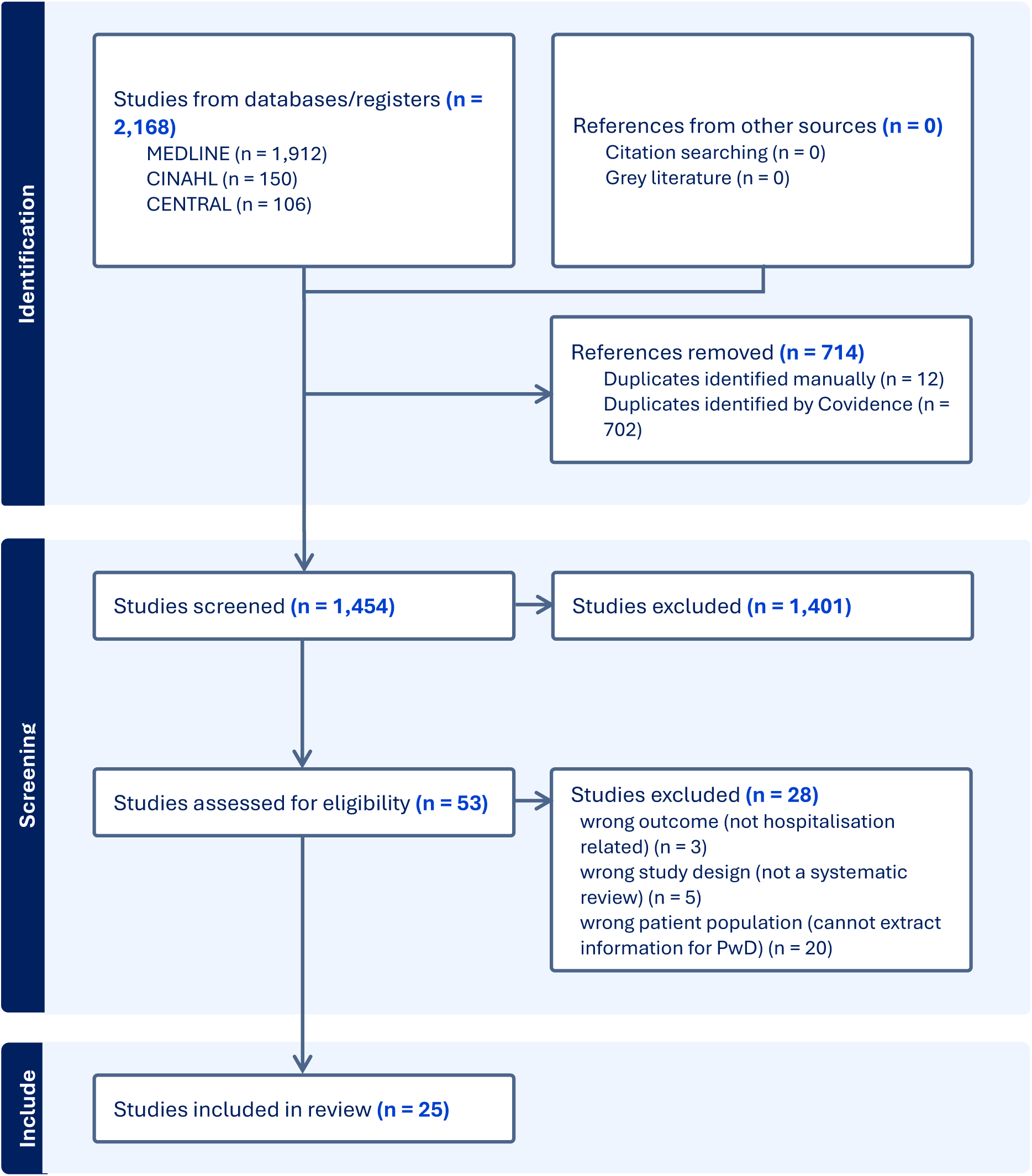
PRISMA diagram.

Seventy-seven unique studies with 1,483,077 participants (RCTs = 47, NRSIs = 30) were included. Twenty-two studies (28.6%) appeared in multiple reviews (Appendix Figure 1 for citation matrix showing overlap). Most studies were conducted in Europe (n=38), followed by North America (n=29), Australia (n=5), East Asia (n=4), and Southeast Asia (n=1).

Twelve of 25 reviews (48%) assessed multiple intervention types. There was a range of intervention types, and we grouped interventions into broad categories to allow for efficient synthesis of findings. Most of the reviews examined the effects of case management (n=15) on hospitalisation of people with dementia, followed by multidisciplinary team management (MDT) (n=9), physical activity interventions (n=4), education/ training programmes (n=4), type of hospital setting (n=3), group activities (n=3), post-operative/ discharge interventions (n=2), palliative care (n=2), counselling (n=2), volunteering (n=2), carer interventions (n=1), and advance care planning (n=1). Studies and the definitions of interventions they included are summarised in Table 1.

**Table 1:**
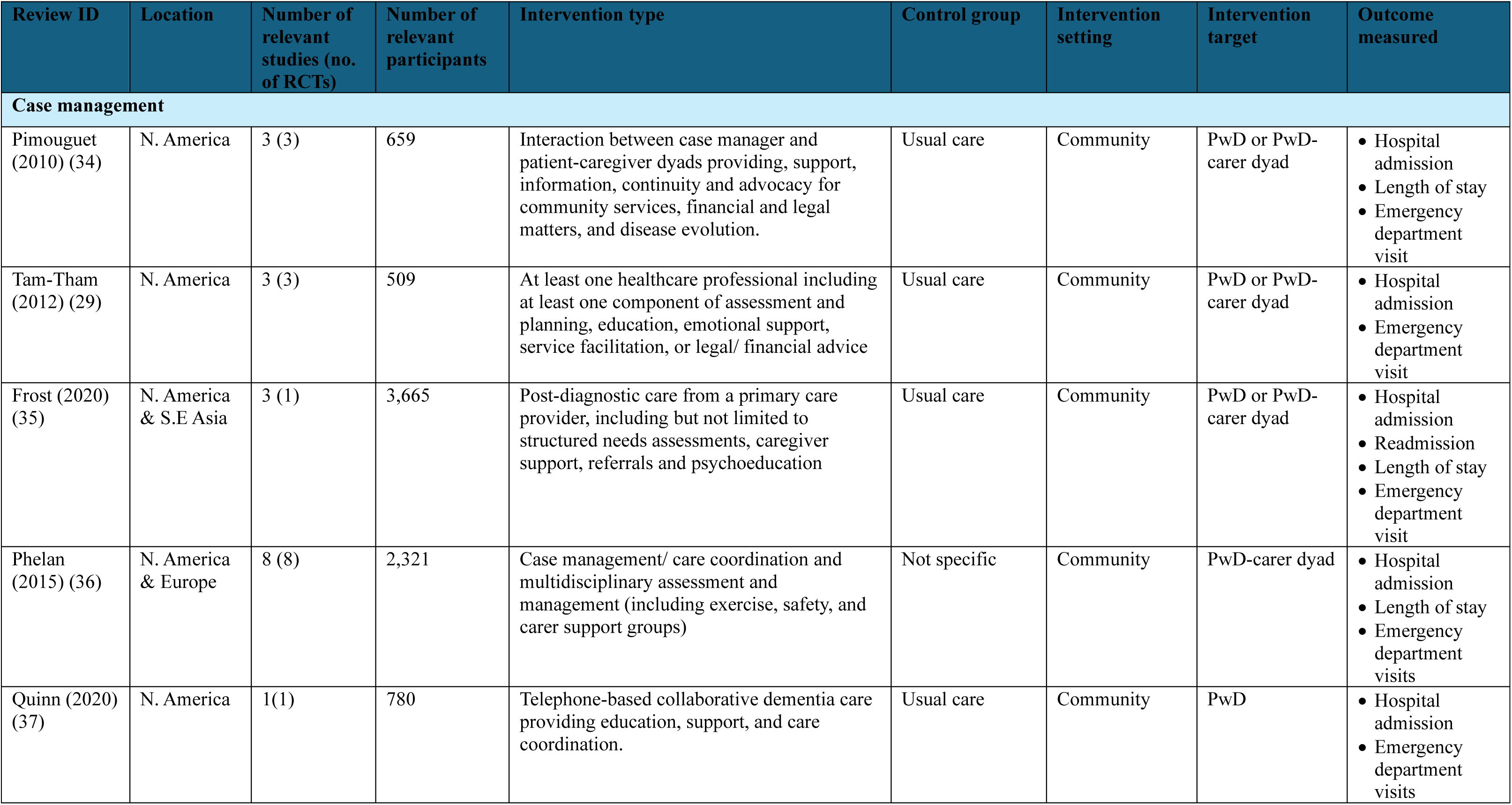
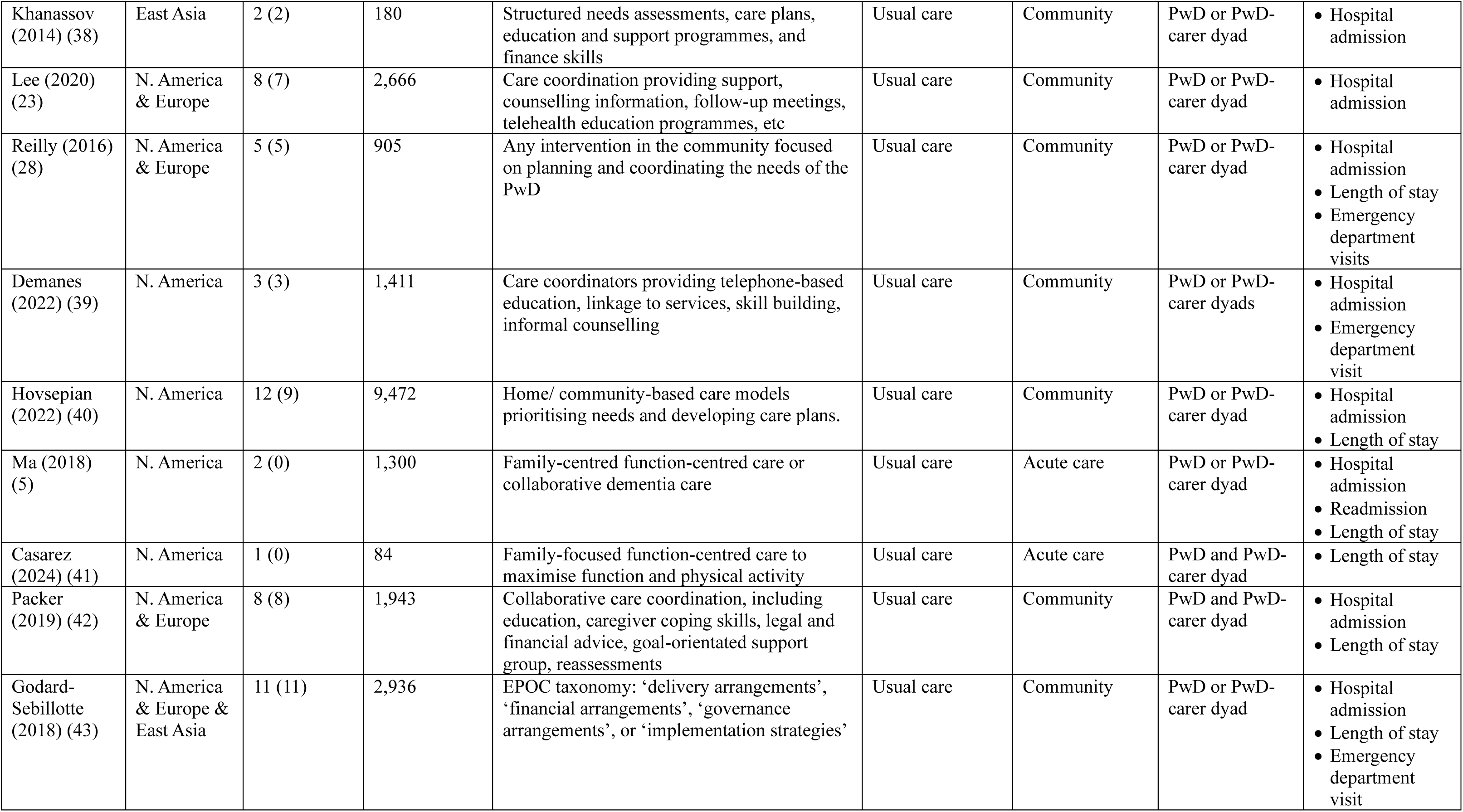
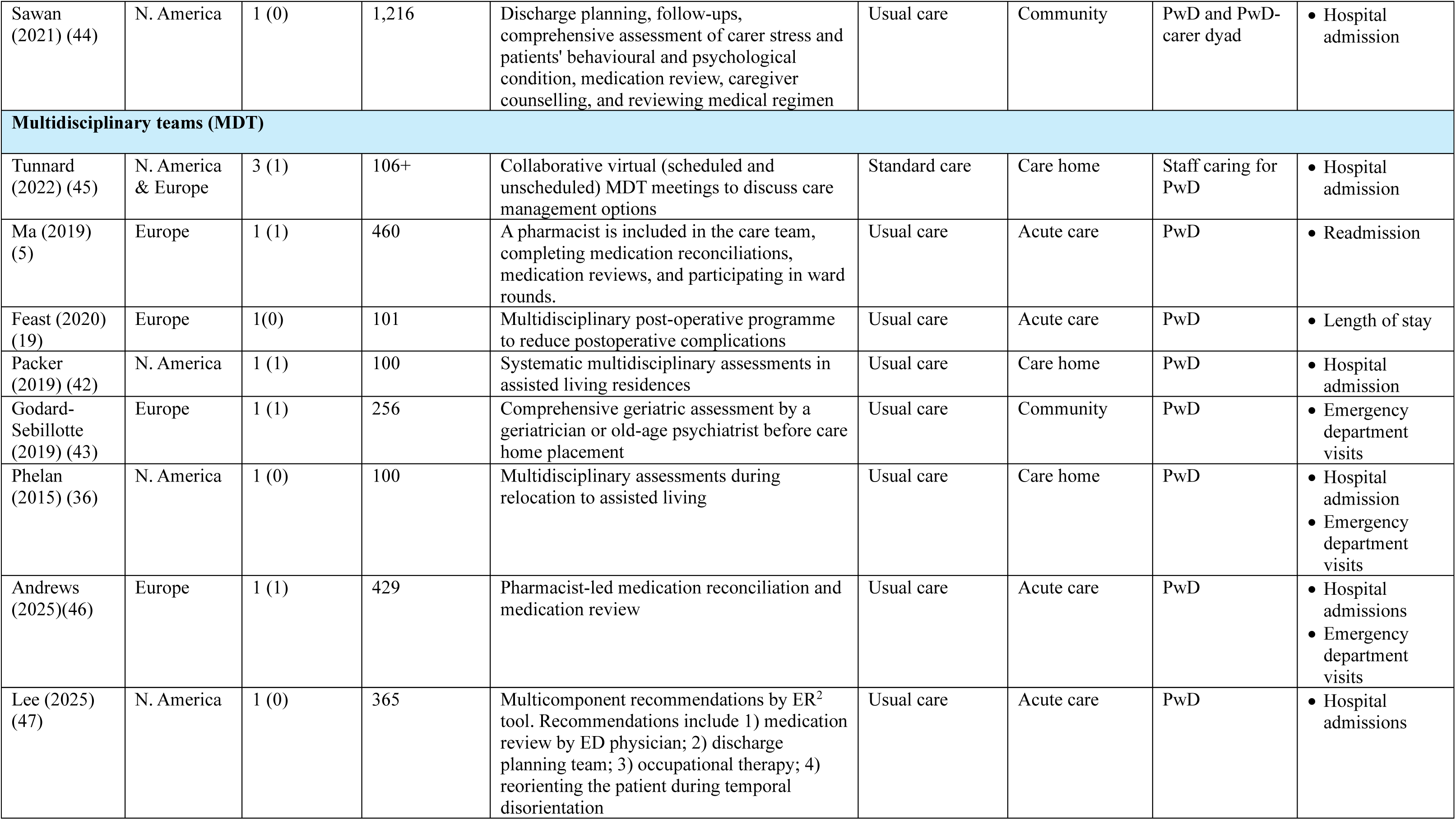
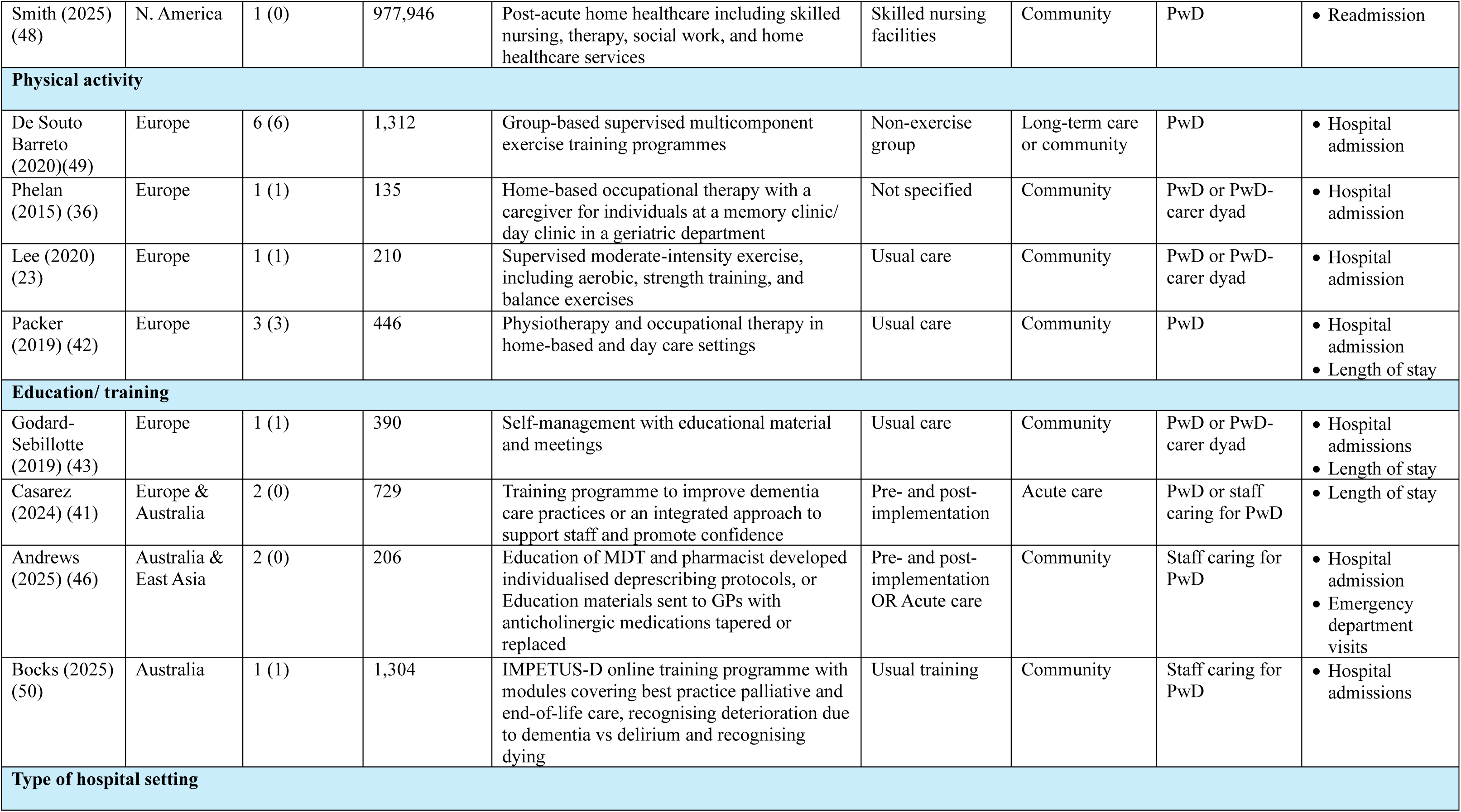
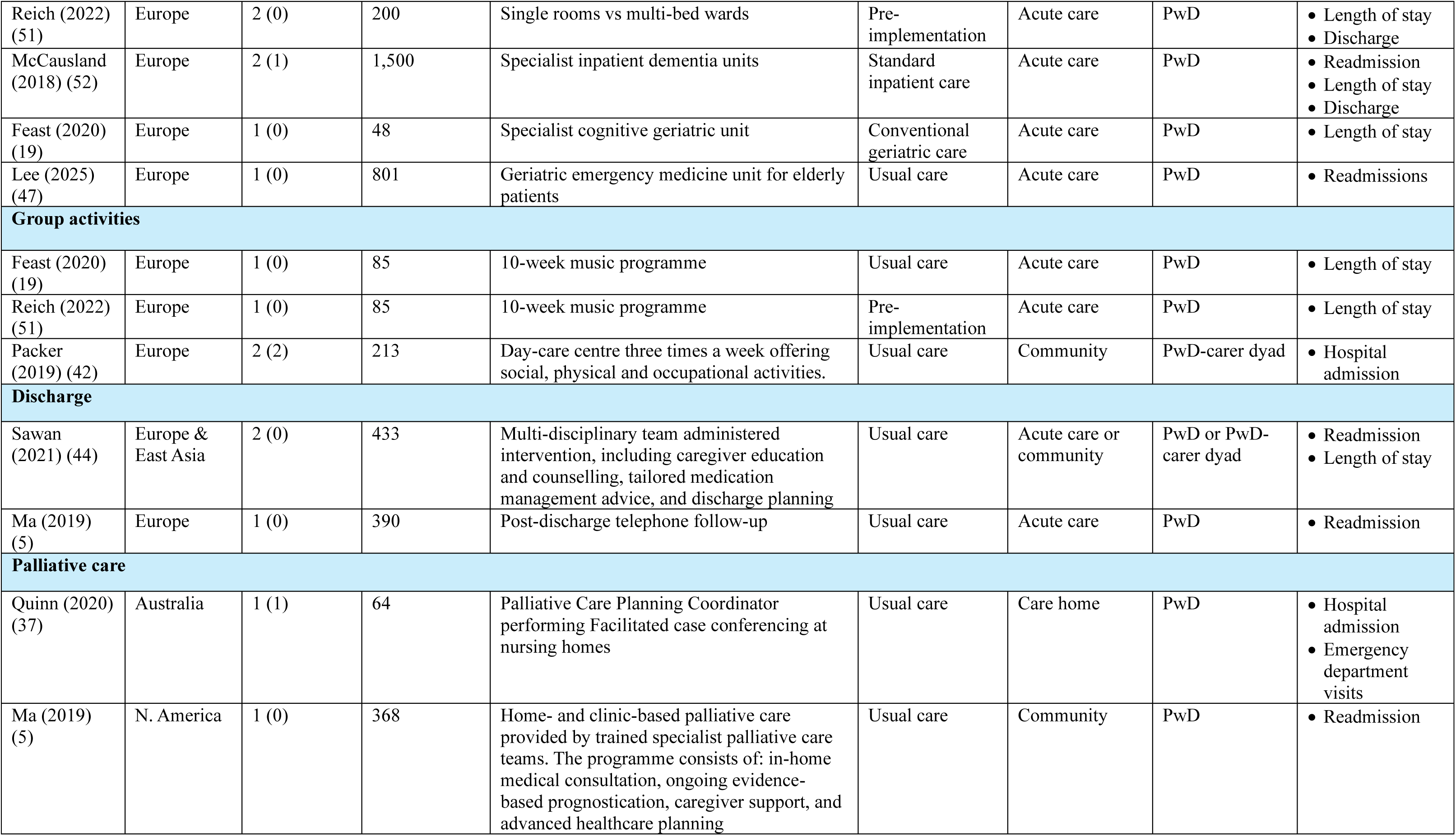
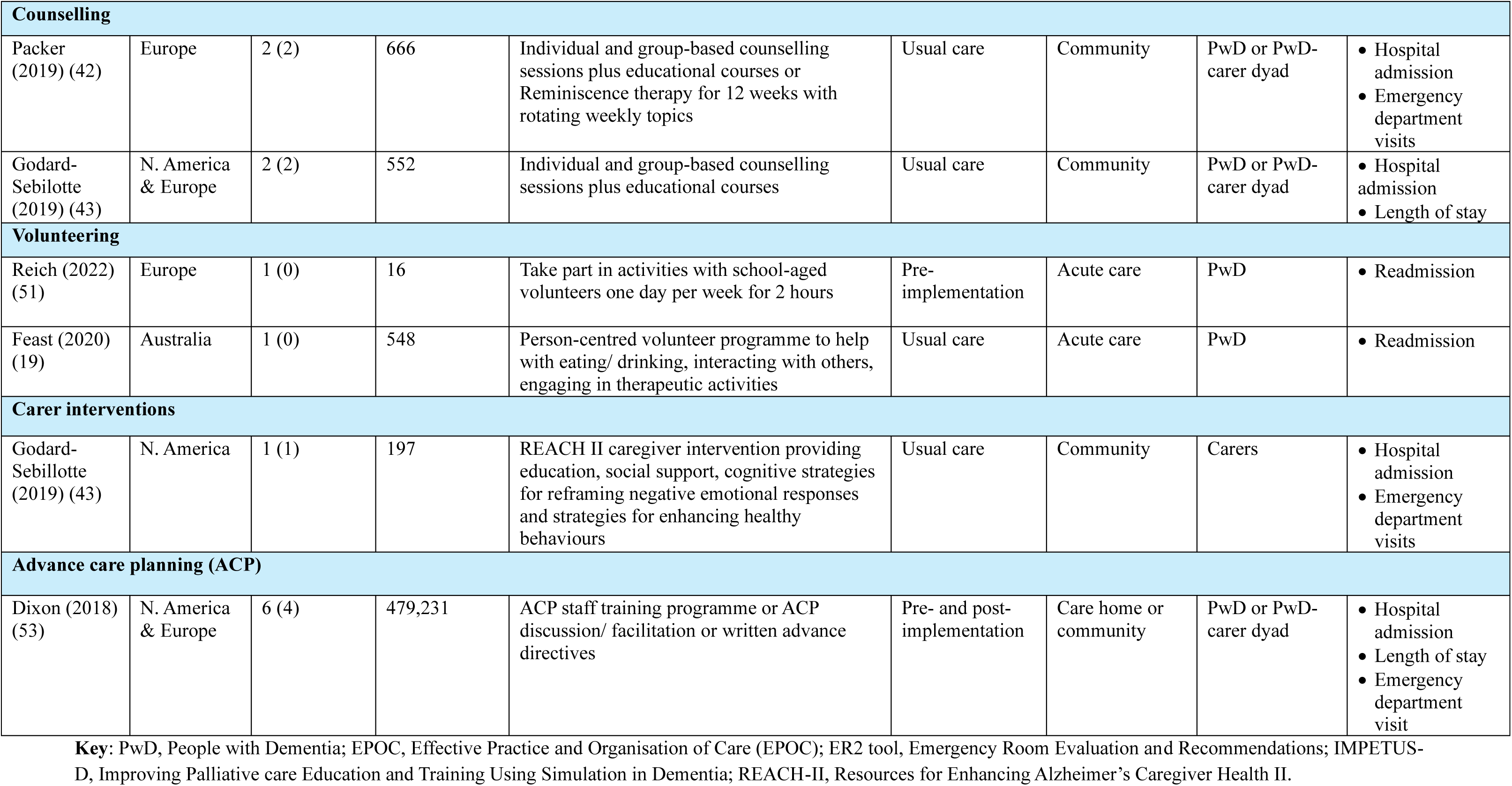
Descriptive table of systematic reviews including the types of studies reviewed, setting and outcome.

According to AMSTAR II criteria (Appendix Table 2), three reviews were high quality (19, 28, 54), eight were moderate quality (29, 37, 40, 41, 45–47, 52), four were low quality (23, 38, 43, 51) and ten were critically low quality (5, 34–36, 39, 42, 44, 50, 53, 55). Issues leading to risk of bias and downgrading of quality mainly related to study protocols not being pre-specified, search strategies that were insufficiently comprehensive, and inadequate methods for assessing risk of bias.

**Table 2:** Summary of evidence and quality of findings for the effect of case management interventions on hospitalisation in people with dementia.

| Quality | Number of studies (of which RCTs) | Sample size | Intervention subtype | Results |
| --- | --- | --- | --- | --- |
| **High** | 5 (5) | 905 | Case management | A meta-analysis found no significant advantage in case management group for reducing risk of hospital admission (OR 0.87 [95% CI: 0.59,1.3]) (28)<br>A smaller meta-analysis (28) on LoS associated with hospital admission, found a small but significant reduction in the control group at 6 months (MD 0.63 [95% CI: 0.40,0.86]; I<sup>2</sup>=40%) (59-61)<br>One small study revealed a significant reduction in the number of days in hospital units or residential homes in the case management group at 6 (MD -5.8 [95% CI: -7.93, -3.67]) and 12 months (MD -7.7 [95% CI: -9.38, -6.02]) (58) |
| **Moderate** | 15 (13) | 7,544 | Case management | One pooled estimate found no significant risk of hospitalisation in case management group (RR 1.00 [95% CI: 0.76, 1.33]; p=0.984) (29)<br>Thirteen studies found no significant difference between groups for hospital admissions (62); (RR 1.12 [95% CI 0.87, 1.68]) (63); (RR 0.96 [95% CI 0.69, 1.34])(64); (RR 1.20 [95% CI 0.64, 2.25]) (60); (RR 0.93 [95% CI 0.69, 1.24]) (71); (RR 1.37 [95% CI 0.38, 4.94])(66); (RR 1.16 [95% CI 0.67-1.99]) (59); (RR 0.57 [95% CI 0.27, 1.22]) (58); (RR 0.88 [95% CI 0.35, 2.21])(67); (RR 0.46 [95% CI 0.23, 0.92])(68); (RR 1.21 [95% CI 0.87, 1.68])(69); (RR 1.04 [95% CI 0.93, 1.17])(70); (MD -0.08 [95% CI -0.29,0.13]) (65); (RR 1.86 [95% CI 0.84, 4.09])(72)<br>Unpublished data found a significant reduction in hospitalisation (RR 0.46 [0.23,0.92]) and LoS (MD -0.34 [95% CI: -0.57, 0.12])(74)<br>Seven studies found no significant difference between groups for ED visits (73) (RR 0.73 [95% CI 0.32, 1.63]) (58); (RR 0.80 [95% CI 0.35, 1.84]) (67); (RR 1.35 [95% CI 0.85, 2.15]) (68); (RR 1.06 [95% CI 0.78, 1.44])(69); (MD -0.17 [95% CI -0.56, 0.22])(65); (RR 0.84 [95% CI 0.49, 1.44]) (72)<br>Seven studies found no significant difference between groups for LoS (MD 0.03 [95% CI -0.25-0.27])(63); (MD -0.16 [95% CI -0.39, 0.07])(64); (MD -0.15 [95% CI -0.51, 0.20]) (59); (MD -2.10 [95% CI -6.9, 2.70]) (58); (MD -1.5 [95% CI -6.14, 3.14]) (67); (MD 0.16 [95% CI -1.69, 1.91])(69); (MD 0.06 [95% CI -0.10, 0.22]) (70).<br>One study found greater LoS in case management group (RR 3.80 [95% CI 3.07, 4.53]) (72) but one study found a small reduction in LoS for case management group (MD -0.37 [-0.66, -0.08])(68) |
|  | 2 (2) | 938 | Virtual case management | No significant effect of intervention on hospital admissions (75, 80) (6 months ($\beta$ 0.09 [95% CI: -0.05, 0.2]) 12 months ($\beta$ -0.03; [95% CI: -0.18, 0.12]) (75) |
| **Low** | 16 (13) | 7,027 | Case management | Eight studies reported no significant differences between groups for hospital admissions (60, 62-66, 69, 76), specifically (ES 1.39 [95% CI 0.39, 4.92])(66); (ES 1.26 [95% CI 0.58, 2.75])(60) (CM 13.5% vs Control 15.9%; $p=0.74$) (64); (DID -1 [95% CI -13,11]) (62)<br>A meta-analysis found no effect for multifactorial interventions and treatments on hospital admissions (RR 1.29 [95% CI 0.67, 2.51])(23). Sensitivity analysis also found no significant effect (RR 0.82 [95%CI 0.36, 1.89])(23)<br>Three studies found reductions in hospital admissions (43% reduction) (79); (14 vs 26%) (78) (6.7% to 0.8% after 6 months for ACSC hospital admissions) (77)<br>Two studies found evidence of reduced ED visits in the intervention groups compared to controls (28% VS 49%)(78) (25% VS 43%) (79)<br>Three studies found reduced LoS for PwD in intervention groups compared to controls ($d=1.9$; unpublished data) (58) ($d=0.224$; unpublished data) (67); (5 vs 7 days) (78).<br>For PwD with at least one hospital admission, severe cognitive impairment patients had fewer admissions at 6 months compared to milder symptoms ($\beta$ -0.29; $p=0.01$) (71) ($\beta=-3.49$; $p<0.10$)(76) |
|  | 3 (3) | 4,370 | Virtual case management | No significant effect of intervention on hospital admissions (75, 80, 81) (6 months ($\beta$ 0.09 [95% CI -0.05, 0.2]) 12 months ($\beta$ -0.03 [95% CI -0.18, 0.12]) (75) (2 year hospitalisations; 31.8% v 34.7%; OR 0.88; $p=0.23$)(81) |
| **Critically low** | 2 (0) | 1,300 | Fam-FFC | No significant difference in LoS (4.0 (SD 2.1)) compared to control (4.4 (SD 2.0)).<br>PwD receiving fam-FFC had significantly lower risk of 30-day readmission ($X^2=5.8$; $p=0.02$) (82) |
|  |  |  | Collaborative dementia care | PwD receiving collaborative dementia care had lower rates of readmission (11% vs 20%), ED visits (28% vs 49%), and hospital admission (14% vs 26%) compared to control (78) |

**Key:** PwD, people with dementia; LoS, length of stay; ED, emergency department; fam-FFC, family-focused function-centred; $\beta$ , beta coefficient; OR, odds ratio; CI, confidence interval; RR, risk ratio; MD, mean difference.

**Table 3:**
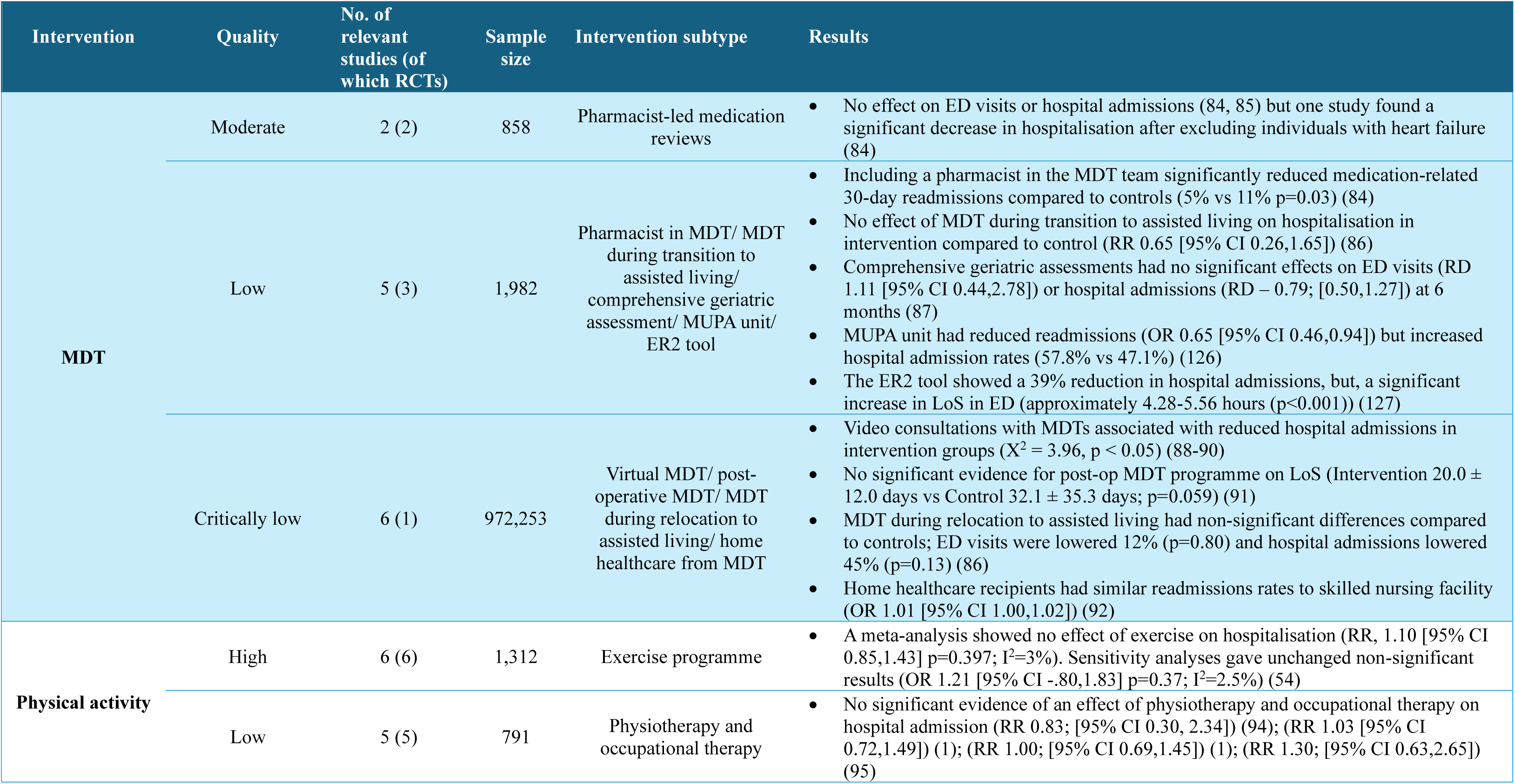
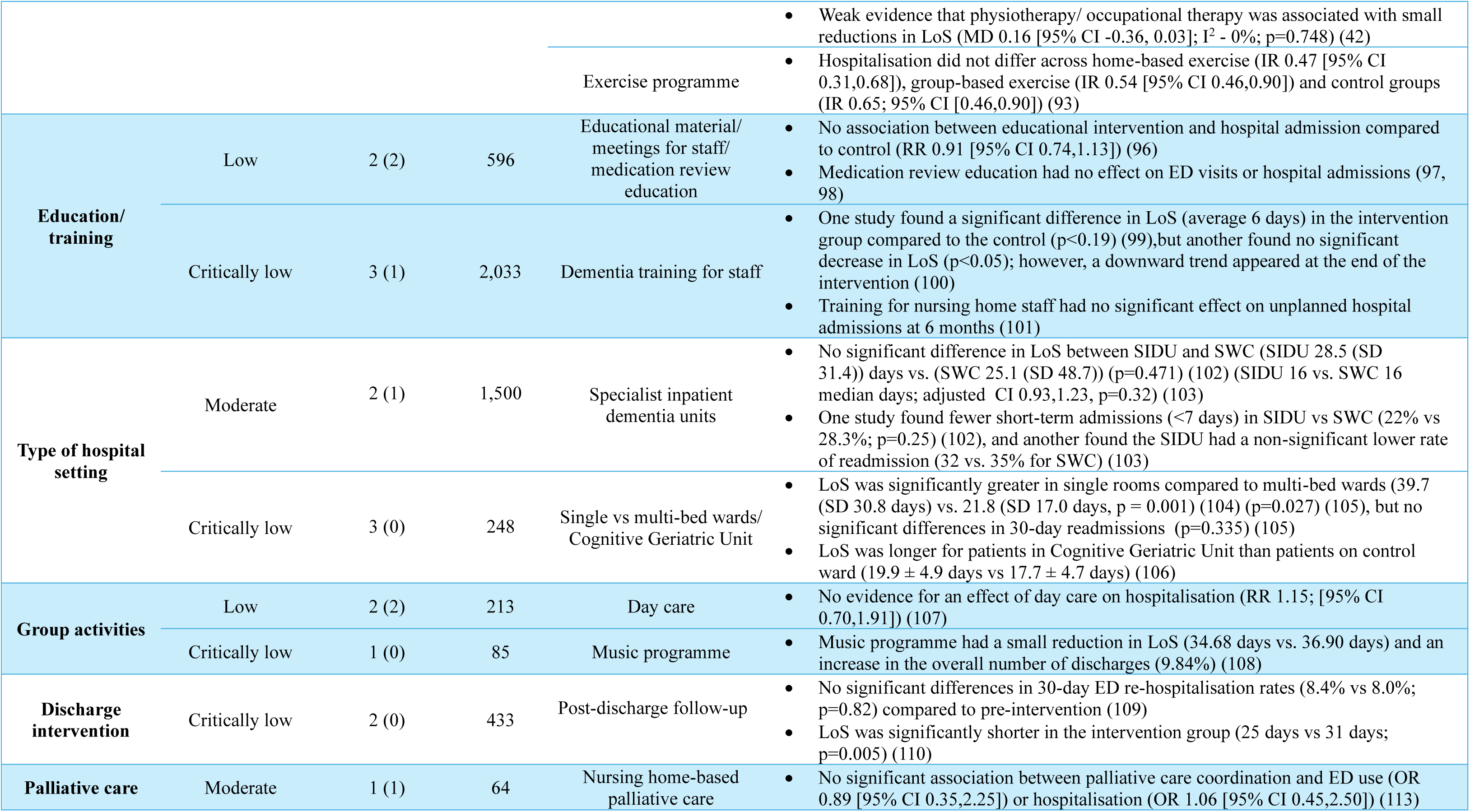
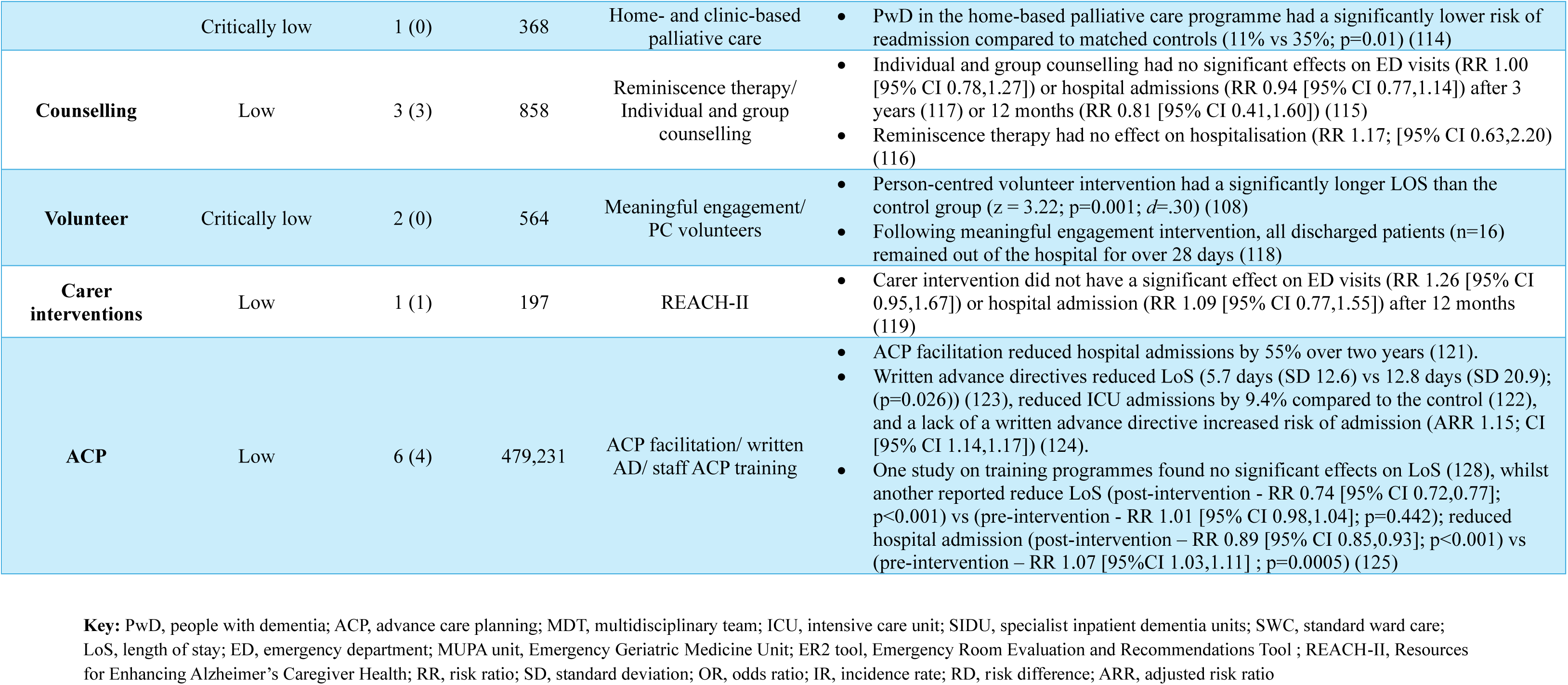
Summary of evidence and quality of findings for the effect of psychosocial interventions on hospitalisation in people with dementia.

The GRADE rating for each intervention within each review (Appendix Table 1) showed high certainty of evidence for findings from two reviews on case management and physical activity. All 11 primary studies in these reviews were RCTs with 905 participants receiving case management and 1,312 participants in the exercise programme. Seven reviews provided moderate certainty of evidence, and these evaluated case management, palliative care, multidisciplinary teams and the type of hospital setting interventions, in 24 primary studies (21 RCTs) with a pooled sample of 10,904 participants. Twelve reviews had low certainty of evidence, and evaluated case management, physical activity, multidisciplinary teams (MDT), advance care planning (ACP), education/ training, group activities, counselling, and carer interventions in 40 primary studies (33 RCTs) with 495,490 participants. Eight reviews had critically low certainty, and evaluated case management, palliative care, type of hospital setting, MDT, education/ training, discharge, group activities, and volunteering in 19 primary studies (1 RCT) with 975,980 participants. Common reasons for downgrading evidence GRADE were issues with risk of bias and imprecision.

### Case management

Based on previous definitions (38, 56, 57), case management was categorised as an intervention with a clinical lead (case manager) responsible for advocating for, planning, and facilitating healthcare services to support the individual and family members care needs to promote high quality outcomes; remote/virtual delivery of case management was assessed in some studies.

#### High quality

One review of five studies examining the effects of case management interventions reported no significant reduction in hospital admissions (28). One study in this review found a significant decrease in the number of days in a hospital unit or residential home in the case management group at 6 (MD -5.8 [95% CI: -7.93, -3.67]) and 12 months (MD -7.7 [95% CI: -9.38, -6.02]) although it was not possible to differentiate hospital and residential home stays (58). A meta-analysis of three studies reported shorter length of stay in the control group compared to intervention group (MD 0.63; [95% CI: 0.40,0.86]; n = 341; I^2^=40%; p<0.001) (59–61).

#### Moderate quality

##### Case management

Thirteen studies reported no significant differences between groups for hospital admission (58–60, 62–72), ten studies found no significant difference between groups for ED visits (58, 65, 67–69, 72, 73), and six studies found no significant difference between groups for LoS (58, 59, 63, 64, 67, 69, 70). However, one review included unpublished data from an RCT which reported a significant reduction in hospital admissions (RR 0.46 [95% CI 0.23,0.92]) and length of stay (MD -0.34 [95% CI: -0.57, 0.12]) in the case management group compared to controls (74). Moreover, one study found greater LoS in the case management group (RR 3.80 [95% CI 3.07, 4.53]) (72) whilst another found a small reduction in LoS for case management group (MD -0.37 [-0.66, -0.08])(68)

##### Virtual case management

One RCT reported no significant effect of case management intervention on hospital admissions at either 6 or 12 months compared to the control group (75).

#### Low quality

##### Case management

Eight studies found no significant differences between groups for hospital admissions (60, 62–66, 69, 76) and a small meta-analysis found no effect of multifactorial interventions and treatments on hospital admissions (RR 1.29 [95% CI 0.67, 2.51])(23) even after making adjustments in sensitivity analyses.

Hospital admissions for ambulatory care-sensitive conditions decreased in the case management group compared to controls after 6 months (6,7% to 0.8%) (77). The Healthy Aging Brain Centre reported reductions in ED visits (28% vs 49%), hospital admissions (14% vs 26%), and length of stay (5 days vs 7 days) compared to controls (78) Another study also found a reduction in ED visits compared to the control group (25% vs 43%) (79) and unpublished data from two additional studies demonstrated weak effects on reduced length of stay (58, 67). Moreover, amongst PwD with at least one hospital admission, those with severe cognitive impairment had fewer hospital admissions after 6 months compared to those with milder cognitive impairment (*β* -0.29; p=0.01) (71) (*β* = -3.49; p<0.10)(76).

##### Virtual case management

Three studies reported no significant effect of technology-enabled case management on hospital admissions at 6-, 12-, or 24-month follow-ups (75, 80, 81).

#### Critically low quality

##### Family-focused function-centred care (fam-FFC)

One study reported no significant difference in length of stay between the fam-FFC group and controls. However, people with dementia receiving fam-FFC had a significantly reduced risk of 30-day readmissions (χ^2^ = 5.8; p=0.02) (82).

##### Collaborative dementia care

One study reported that people with dementia receiving collaborative dementia care had lower rates of readmission (11 vs 20%), ED visits (28 vs 49%), and hospital admissions (14 vs 26%) compared to controls (78).

##### Summary

There was high quality evidence to suggest that case management is not effective in reducing hospital admissions or length of stay in people with dementia and moderate and low certainty evidence suggesting virtual case management is also ineffective.

### Multidisciplinary (MDT) teams

Included MDT interventions varied in structure and composition, but these were often characterised by multidisciplinary services from various healthcare providers, including, neurologists, geriatricians, social workers, nurse practitioners and physical/ occupational therapists (83).

#### Moderate quality

Two RCTs (n=858) found no effect of pharmacist-led medication reviews on hospital admissions or ED visits (84, 85) but one study found a significant decrease in hospitalisation after adjusting for heart failure (84).

#### Low quality

One RCT (n=460) found that including a clinical pharmacist in the hospital MDT team produced significantly lower medication-related 30-day readmissions compared to control (5% vs 11%) (84). However, comprehensive geriatric assessments and MDT assistance during transition to assisted living did not effect hospitalisation (86, 87).

#### Critically low quality

Video consultations with MDT showed significant evidence of reduced hospital admissions (X^2^=3.96; p<0.05) (88–90). However, post-operative MDT interventions, MDT during relocation to assisted living, and home healthcare groups found no significant reductions in hospitalisation (86, 91, 92).

##### Summary

Moderate quality evidence suggests pharmacist-led medication reviews can reduce hospitalisation in dementia only after adjusting for physical comorbidities.

### Physical activity

Physical activity interventions included an intervention focused on improving strength, mobility and balance through exercise groups or physical therapy programmes.

#### High quality

One review of six RCTs examining the effects of supervised exercise programmes at home found no significant impact on hospitalisations compared controls (RR, 1.10 [95% CI 0.85-1.43]), even after conducting sensitivity analysis (OR 1.12 [95% CI: 0.90,1.83]; p=0.37) (49).

#### Low quality

One RCT (n=210) found hospitalisation did not differ across home-based exercise, group-based exercise, or control groups (93). Four RCTs (n=581) found there were also no significant reductions in hospital admissions after physiotherapy and occupational therapy interventions (84, 94, 95), but there was weak evidence that physiotherapy and occupational therapy were associated with small reductions in length of stay (MD 0.16 [95% CI -0.36, 0.03]) (95).

##### Summary

There is high quality evidence that while in general supervised exercise programmes for people with dementia may be helpful in other domains they do not decrease the likelihood of hospitalisations.

### Education/ training

Interventions were categorised as education/ training programmes provided they aimed to improve staff confidence, knowledge or understanding of dementia care through face-to-face training, collaborative meetings, or reading materials.

#### Low quality

Two RCTs (n=596) assessing educational materials, meetings with healthcare professionals produced, and medication review education, found no significant association with hospital admissions compared to controls (96–98).

#### Critically low quality

Whilst one NRSI (n=68) reported dementia training for staff produced a significant reduction in length of stay (average 6 days) (99), another NRSI (n=661) reported no significant difference in length of stay, but a downward trend was established towards the end of the intervention (100). An RCT (n=1304) found no effect for nursing home staff training on hospital admissions at 6 months (101)

##### Summary

There is no good evidence of an effect of education or training but a lack of evidence

### Type of hospital setting

Type of hospital setting interventions focused on situational variables in acute or community care that differed to regular care settings or were tailored to people with dementia.

#### Moderate quality

Two studies (n=1500) found no significant difference reported between specialist inpatient dementia units and control groups for length of stay (SIDU 28.5 (SD 31.4)) days vs. (SWC 25.1 (SD 48.7))(102) (SIDU 16 vs SWC 16 median days) (103). Whilst one RCT (n=600) found non-significant reductions in readmission rates (103), another NRSI (n=900) reported fewer short-term admissions in the intervention group compared to controls (22% vs 28.3%) (102).

#### Critically low quality

Two NRSIs (n=200) examining single-bed rooms were associated with significantly reduced length of stay compared to multi-bed wards (39.7 (SD 30.8 days) vs. 21.8 (SD 17.0 days)) (104, 105) but there was no significant difference in 30-day readmissions (105). Cognitive Geriatric Units (NRSI, n=48) were also associated with increased length of stay compared to control (19.9 ± 4.9 vs 17.7 ± 4.7 days) (106).

##### Summary

There was a lack of good quality evidence but to date there is no evidence that the type of hospital setting makes a difference

### Group activities

Group activities referred to social activities based within the community or residential care settings.

#### Low quality

Two RCTs (n=213) found day care interventions had no effect on hospitalisation compared to control (107).

#### Critically low quality

One NRSI (n=85) found music programmes demonstrated a small reduction in length of stay (34.68 days vs 36.90) and an increase in the total number of discharges (9.84%) compared to the control group. (108).

##### Summary

There is no good evidence of an effect of group activities but a lack of evidence

### Discharge intervention

Discharge interventions consisted of follow-ups from healthcare providers, guidance for people with dementia and caregivers, medication management or discharge planning for people with dementia discharged from acute care.

#### Critically low quality

One NRSI (n=390) found no significant differences between post-discharge and control groups for 30-day ED readmissions (8.4% vs 8.0%) (109). Another NRSI (n=43) but one study did find evidence of a significant reduction in length of stay (25 days vs 31 days) (110).

##### Summary

There is no good evidence of an effect of discharge intervention but a lack of evidence

### Palliative care

Based on previous definitions, palliative care interventions were categorised as interdisciplinary patient-centred care focusing on physical, psychosocial and spiritual needs with careful reviews and management to best promote quality of life (111, 112).

#### Moderate quality

One RCT (n=64) reported no significant association between palliative care and ED visits or hospital admissions compared to the control group (113).

#### Critically low quality

One observational study (n=368) reported that people with dementia receiving home-based palliative care had a significantly lower risk of readmissions compared to matched controls (11% vs 35%) (114).

##### Summary

There is no good quality evidence that palliative care reduces hospital admissions or ED visits but a lack of evidence.

### Counselling

Counselling interventions included individual or group-based talking sessions led and managed by healthcare providers

#### Low quality

Three RCTs (n=858) found no significant effect for individual/ group counselling or reminiscence therapy on hospitalisation (115–117).

##### Summary

There is no good evidence for an effect of counselling on hospitalisation but a lack of evidence

### Volunteers

Volunteer programmes were categorised as interventions focusing on social connections and emotional support between volunteers and people with dementia or caregivers through activities or discussions to enhance quality of life.

#### Critically low

All patients (n=16) discharged following a meaningful engagement intervention remained out of the hospital for over 28 days (108). Also, the person-centred volunteer intervention had a significantly longer length of stay than the control group (z = 3.22; p=0.001; *d*=.30) (118).

##### Summary

There is no good evidence of an effect of volunteers but a lack of evidence

### Carer intervention

Carer interventions refer to individualised psychosocial support for caregivers of people with dementia including but not limited to social support, stress management, or skills training.

#### Low quality

One RCT (n=197) found carer interventions did not have a significant effect on ED visits or hospital admissions after 12 months (119).

##### Summary

There is no good evidence of an effect of carer interventions but a lack of evidence

### Advance Care Plans (ACP)

Advance care planning aims to offer the opportunity to have meaningful discussions planning future care and support, including medical treatment and end-of-life support, whilst people have the mental capacity to do so (120).

#### Low quality

In one review of six studies (four RCTs) and 479,231 participants, ACP facilitation was found to reduce hospital admissions by 55% over 2 years (121), and written directives found to reduce ICU admissions (9.4%) and length of stay (5.7 days (SD 12.6) vs 12.8 days (SD 20.9)) (122, 123). Also, the absence of an advance directive was found to increase the risk of hospital admission (ARR 1.15; CI [95% CI 1.14,1.17]) (124). However, training programmes for delivering ACP found mixed effects on the length of stay compared to control (125).

##### Summary

There was only low-quality evidence of ACP and written directives, but results were promising, suggesting they reduced hospital admission. Training staff to increase delivery of ACP had mixed results.

## Discussion

Our umbrella review is the first to examine the effects of psychosocial and healthcare interventions on reducing hospitalisation in PwD. We found 25 systematic reviews, including 77 unique studies (RCTs = 47; NRSIs = 30) with 1,483,077 participants. Overall, there was high certainty that case management and exercise programmes had no significant effect on hospital admissions or length of stay. The most promising interventions were ACP facilitation which resulted in significantly reduced hospitalisation; and including clinical pharmacists in MDT interventions which significantly reduced medication-related readmissions but both had low/moderate certainty of evidence for efficacy. There was moderate-certainty evidence that palliative care did not reduce hospitalisation. Other interventions including education, group activities, counselling, and carer interventions were not shown to affect hospitalisation but there was low certainty for this evidence.

### Strengths and limitations

We conducted a comprehensive search and employed a rigorous systematic approach to data collection and synthesis. Nonetheless, most evidence was from high-income countries, meaning we cannot generalise our findings to low- and middle-income countries or healthcare systems. In addition, there is considerable heterogeneity across interventions and outcomes, meaning our review was unable to meta-analyse to enhance statistical power or categorise results definitively by intervention and outcome. By their nature, umbrella reviews omit recent primary studies of potential value so although we updated our searches, our review cannot include the full scope of primary research. Whilst we considered confounders during GRADE assessments, we were unable to establish the influence of confounders on the association between interventions and hospitalisation in PwD in NSRIs. Additionally, several reviews included unpublished data obtained through personal correspondence and despite contacting authors for clarification, we cannot validate this data and results. Finally, little research evidence was of high certainty, meaning we cannot establish the true effect of each intervention on hospitalisation in dementia without further robust and high-quality RCTs.

### Interpretation in the context of other research

The most studied intervention was case management, and though it might be expected that the presence of a clinical lead (case manager) responsible for advocating for, planning, and facilitating access to healthcare services to support the individual and family would be effective, we did not find this. A recent scoping review found case management for PwD is often poorly defined and described (129), which may explain the negative results. Further, distrust among healthcare professionals and barriers to accessing health and social care data hinders information sharing, working relationships and case management efficiency (130).

Our findings of reduced hospitalisation following ACP facilitation and written directives are consistent with a previous review (131). Training programmes were not effective possibly because of varied delivery setting or funding, as staff turnover, staff shortages, and organisational culture play a critical role in successful implementation of ACP (132).

Previous systematic reviews assessing exercise effects on hospitalisation in older adults found a reduction in the number of falls following exercise interventions (54, 133). However, in our analysis of studies of PwD, we found strong evidence that these programmes did not reduce hospitalisation suggesting that there may be additional components required beyond general exercise programmes to reduce hospitalisation in PwD. Studies of general older adult populations had similarly found that palliative care had a significant effect on hospitalisation (134, 135), but we found with moderate certainty that there was no effect in PwD, suggesting that palliative care need to be specifically tailored for PwD.

The low certainty of evidence examining the effects of MDT on hospitalisation in dementia is related to small studies lacking sufficient statistical power. However, one review stated that included primary studies only measured ‘acute hospital use’, meaning potentially avoidable admissions might have been excluded (43), which may explain the absence of an observed association. Previous research on the impact of specialist wards on hospitalisation in dementia indicated a potential reduction in readmission (136) and non-significant reductions in length of stay (137). Our results are in line with this but there is insufficient evidence for firm conclusions.

As the evidence assessing the remaining psychosocial interventions is of critically low quality, we are unable to make any recommendations based on the literature. Consequently, further high-quality RCTs implementing standardised interventions and outcomes is required to determine their efficacy in reducing hospitalisation in PwD.

## Conclusion and recommendations

This umbrella review has shown that current evidence for psychosocial and healthcare interventions in reducing hospitalisation in dementia is insufficient to make definitive guidelines for commissioners and policy makers. Clinicians may however consider ACP or written directives as effective interventions to potentially reduce avoidable admissions. Including pharmacists in MDT interventions may also help reduce hospitalisation in dementia care. However, clinicians should acknowledge the high certainty evidence suggesting case management and exercise programmes are ineffective in reducing hospitalisations in PwD. Future robust RCTs are required to improve the certainty of positive recommendations. Specifically, researchers should focus on interventions that show promise but lack sufficient high-quality evidence to support them, such as ACP, MDT interventions, and discharge interventions. Moreover, detailed descriptions of interventions and outcomes using standardised checklists, such as TIDieR, are needed to reduce heterogeneity and determine the true efficacy of each intervention component. Researchers should also explore the efficacy of these interventions across a broad range of countries and their healthcare systems to establish effectiveness in more diverse settings.

### What is already known on this topic

- People with dementia (PwD) are at an increased risk of hospitalisation, stay in hospital for longer and are at higher risk of readmission compared to individuals without dementia
- Hospital admissions can have significant negative effects on PwD, including increased mortality and frailty, more inpatient complications and further cognitive and functional decline
- Current evidence on the effects of psychosocial and healthcare interventions on reducing hospitalisation in dementia is spread across various intervention types and lacks clear efficacy

### What this study adds

- There is high certainty evidence that case management and exercise programmes have no effect on hospitalisation in dementia
- Advance care planning and clinical pharmacists in multidisciplinary teams show promise in reducing hospitalisation in dementia
- Evidence for other psychosocial and healthcare interventions on hospitalisation in PwD is insufficient and of low quality to make strong recommendations

## Ethical Approval

Not needed

## Data availability statement

All data produced for this study is available in the manuscript and appendices.

## Acknowledgements

We would like to acknowledge the contributions of our panel of patient and public involvement representatives.

## Transparency statement

The lead author affirms that the manuscript is an honest, accurate and transparent account of the study being report; that no important aspects of the study have been omitted; and that any discrepancies from the study as originally planned and registered have been explained.

## Contributors

CH was involved in literature searching, data curation, data extraction, data synthesis, quality assessment, and writing the original draft of the manuscript. AF-S was involved in quality assessment, and editing. NA was involved in data curation and data extraction. GL was involved in conceptualisation, data synthesis, supervision and editing. AS was involved in conceptualisation, quality assessment, data curation, data synthesis, supervision, and editing. SG and CR were involved as PPI collaborators and in editing. DD, CG, MH, NM, KW were involved in reviewing and editing. All authors reviewed and approved the final version of the manuscript. Corresponding author (AS) attests that all listed authors meet authorship criteria and that none meeting the criteria have been omitted.

## Copyright

“The Corresponding Author has the right to grant on behalf of all authors and does grant on behalf of all authors, a worldwide licence to the Publishers and its licensees in perpetuity, in all forms, formats and media (whether known now or created in the future), to i) publish, reproduce, distribute, display and store the Contribution, ii) translate the Contribution into other languages, create adaptations, reprints, include within collections and create summaries, extracts and/or, abstracts of the Contribution, iii) create any other derivative work(s) based on the Contribution, iv) to exploit all subsidiary rights in the Contribution, v) the inclusion of electronic links from the Contribution to third party material where-ever it may be located; and, vi) licence any third party to do any or all of the above.”

## Appendix

### Search strategy

1. Meta-analysis as Topic/
2. meta analy$.tw.
3. metanaly$.tw.
4. Meta-Analysis/
5. (systematic adj (review$1 or overview$s1)).tw.
6. exp Review Literature as Topic/
7. or/1-6
8. cochrane.ab.
9. embase.ab.
10. (psych lit or psyclit).ab.
11. (psychinfo or psycinfo).ab.
12. (cinahl or cinhal).ab.
13. science citation index.ab.
14. bids.ab.
15. cancerlit.ab.
16. or/8-15
17. reference list$.ab.
18. bibliograph$.ab.
19. hand-search$.ab.
20. relevant journals.ab.
21. manual search$.ab.
22. or/17-21
23. selection criteria.ab.
24. data extraction.ab.
25. 23 or 24
26. Review/
27. 25 and 26
28. Comment/
29. Letter/
30. Editorial/
31. animal/
32. human/
33. 31 not (31 and 32)
34. or/28-30,33
35. 7 or 16 or 22 or 27
36. 35 not 34
37. (dementia or dement$ or “Alzheimer’s dementia”).tw.
38. Dementia.mp.
39. 37 or 38
40. advance care plan$.mp.
41. case manag$.mp.
42. psychoeduca$.mp.
43. self-manag$.mp.
44. palliative care.mp.
45. advance direct$.tw.
46. Intervention.mp.
47. therapy.mp.
48. evaluat$.mp.
49. train$.mp.
50. rehabilitation.mp.
51. treatment$.mp.
52. or/40-52
53. hospita$.mp.
54. admis$.mp.
55. admit$.mp.
56. readmi$.mp.
57. length of stay.mp.
58. discharg$.mp.
59. Hospitalization/
60. Emergency Room Visits/
61. Or/54-61
62. 36 and 39 and 53 and 62

**Table 1:** Grade approach.

| Author (Year) | Intervention type | No. of studies (of which RCTs) | Downgrading |  |  |  |  | Upgrading |  |  | GRADE quality |
| --- | --- | --- | --- | --- | --- | --- | --- | --- | --- | --- | --- |
|  |  |  | Risk of bias | Inconsistency | Indirectness | Imprecision | Publication Bias | Magnitude of effect | Dose-response gradient | Direction of bias |  |
| **Reich (2022)** | Situational | 2 (0) | ⊖ | ⊕ | ⊕ | ⊖ | N/A | Yes | N/A | No | Critically low |
|  | Group activities | 1 (0) | ⊖ | ⊕ | ⊕ | ⊕ | N/A | No | N/A | No |  |
|  | Volunteer | 1 (0) | ⊖ | ⊕ | ⊕ | ⊖ | N/A | No | N/A | No |  |
| **DeSouto Barreto (2021)** | Physical activity | 6 (6) | ⊕ | ⊕ | ⊕ | ⊕ | ⊕ | No | N/A | No | High |
| **Pimouguet (2010)** | Case management | 3 (3) | ⊖ | ⊕ | ⊕ | ⊖ | N/A | No | N/A | No | Low |
| **Dixon (2018)** | ACP | 6 (4) | ⊖ | ⊕ | ⊕ | ⊕ | ⊖ | No | N/A | No | Low |
| **Tam-Tham (2013)** | Case management | 3 (3) | ⊕ | ⊕ | ⊕ | ⊖ | ⊕ | No | N/A | No | Moderate |
| **Frost (2020)** | Case management | 3 (1) | ⊖ | ⊕ | ⊕ | ⊕ | N/A | No | N/A | No | Moderate |
| **Phelan (2015)** | Physical activity | 1 (1) | ⊖ | ⊕ | ⊕ | ⊖ | N/A | No | N/A | No | Low |
|  | MDT | 1 (0) | ⊖ | ⊕ | ⊕ | ⊖ | N/A | No | N/A | No | Critically low |
|  | Case management | 8 (8) | ⊖ | ⊖ | ⊕ | ⊕ | N/A | No | N/A | No | Low |
| **Quinn (2020)** | Case management | 1 (1) | ⊕ | ⊕ | ⊕ | ⊖ | ⊕ | No | N/A | No | Moderate |
|  | Palliative care | 1 (1) | ⊕ | ⊕ | ⊕ | ⊖ | ⊕ | No | N/A | No |  |
| **Lee (2020)** | Physical activity | 1 (1) | ⊖ | ⊕ | ⊕ | ⊖ | N/A | No | N/A | No | Low |
|  | Case management | 8 (7) | ⊖ | ⊖ | ⊕ | ⊕ | N/A | No | N/A | No |  |
| **Reilly (2015)** | Case management | 5 (5) | ⊕ | ⊕ | ⊕ | ⊕ | ⊕ | No | N/A | No | High |
| **Ma (2019)** | Case management | 2 (0) | ⊖ | ⊕ | ⊕ | ⊕ | N/A | No | N/A | No | Critically low |
|  | Palliative care | 1 (0) | ⊖ | ⊕ | ⊕ | ⊖ | N/A | No | N/A | No |  |
|  | Discharge | 1 (0) | ⊖ | ⊕ | ⊕ | ⊖ | N/A | No | N/A | No |  |
|  | MDT | 1 (1) | ⊖ | ⊕ | ⊕ | ⊕ | N/A | No | N/A | No | Low |
| **Sawan (2021)** | Case management | 1 (0) | ⊖ | ⊕ | ⊕ | ⊕ | N/A | No | N/A | No | Critically low |
|  | Discharge | 2 (0) | ⊖ | ⊕ | ⊕ | ⊖ | N/A | No | N/A | No |  |
| **Demanes (2021)** | Case management | 3 (3) | ⊕ | ⊕ | ⊖ | ⊖ | N/A | No | N/A | No | Low |
| **McCausland (2019)** | Type of hospital setting | 2 (1) | ⊕ | ⊕ | ⊕ | ⊕ | N/A | No | N/A | No | Moderate |
| **Hovsepian (2022)** | Case management | 12 (9) | ⊕ | ⊖ | ⊖ | ⊕ | N/A | No | N/A | No | Low |
| **Tunnard (2022)** | MDT | 3 (1) | ⊕ | ⊕ | ⊕ | ⊖ | ⊕ | No | N/A | No | Critically low |
| **Feast (2020)** | Type of hospital setting | 1 (0) | ⊕ | ⊕ | ⊕ | ⊖ | N/A | No | N/A | No | Critically low |
|  | MDT | 1 (0) | ⊕ | ⊕ | ⊕ |  | N/A | No | N/A | No |  |
|  | Group activities | 1 (0) | ⊕ | ⊕ | ⊕ |  | N/A | No | N/A | No |  |
|  | Volunteer | 1 (0) | ⊕ | ⊕ | ⊕ |  | N/A | No | N/A | No |  |
| **Packer (2019)** | Case management | 8 (8) | ⊖ | ⊕ | ⊕ | ⊕ | N/A | No | N/A | No | Moderate |
|  | Physical activity | 3 (3) | ⊖ | ⊕ | ⊕ | ⊖ | N/A | No | N/A | No | Low |
|  | MDT | 1 (1) | ⊖ | ⊕ | ⊕ | ⊖ | N/A | No | N/A | No |  |
|  | Counselling | 2 (2) | ⊖ | ⊕ | ⊕ | ⊖ | N/A | No | N/A | No |  |
|  | Group activities | 2 (2) | ⊖ | ⊕ | ⊕ | ⊖ | N/A | No | N/A | No |  |
| **Godard-Sebillotte (2019)** | Case management | 11 (11) | ⊖ | ⊕ | ⊕ | ⊕ | ⊕ | No | No | No | Moderate |
|  | Education/ training | 1 (1) | ⊖ | ⊕ | ⊕ | ⊖ | ⊕ | No | No | No | Low |
|  | MDT | 1 (1) | ⊖ | ⊕ | ⊕ | ⊖ | ⊕ | No | No | No |  |
|  | Carer intervention | 1 (1) | ⊖ | ⊕ | ⊕ | ⊖ | ⊕ | No | No | No |  |
|  | Counselling | 2 (2) | ⊖ | ⊕ | ⊕ | ⊖ | ⊕ | No | No | No |  |
| **Casarez (2024)** | Case management | 1 (0) | ⊕ | ⊕ | ⊕ | ⊖ | N/A | No | No | No | Critically low |
|  |  | 2 (0) | ⊕ | ⊕ | ⊕ | ⊖ | N/A | No | No | No |  |
| **Khanassov (2014)** | Case management | 2 (2) | ⊖ | ⊕ | ⊕ | ⊖ | N/A | No | No | No | Low |
| **Andrews (2025)** | Education/ training | 2 (0) | ⊖ | ⊕ | ⊕ | ⊕ | N/A | No | No | No | Critically low |
|  | MDT | 2 (2) | ⊕ | ⊕ | ⊕ | ⊖ | N/A | No | No | No | Moderate |
| **Bocks (2025)** | Education/ training | 1 (1) | ⊖ | ⊕ | ⊕ | ⊕ | N/A | No | No | No | Low |
| **Lee (2025)** | MDT | 2 (0) | ⊕ | ⊕ | ⊕ | ⊕ | N/A | No | No | No | Low |
| **Smith (2025)** | MDT | 1 (0) | ⊖ | ⊕ | ⊕ | ⊕ | N/A | No | No | No | Critically low |

⊕ No concerns
⊖ Serious concerns

**Table 2:** Quality assessment with AMSTAR II tool.

⊕ No concerns ⊖ Serious concerns
| Table 2: Quality assessment with AMSTAR II tool |  |  |  |  |  |  |  |  |  |  |  |  |  |  |  |  |  |  |  |
| --- | --- | --- | --- | --- | --- | --- | --- | --- | --- | --- | --- | --- | --- | --- | --- | --- | --- | --- | --- |
| *Author* | *Question and inclusion* | *Protocol\** | *Study design* | *Search strategy\** | *Study selection* | *Data extraction* | *Exclusion reasons\** | *Inclusion details* | *RoB assessment (RCTs) \** | *RoB assessment (NRSI) \** | *Funding* | *Meta-analysis method (RCTs)\** | *Meta-analysis method (NRSI)\** | *Meta-analysis RoB impact* | *RoB discussion\** | *Heterogeneity discussion* | *Publication bias\** | *Conflict of interest* | *Overall* |
| *Reich* | Yes | Py | Yes | No | Yes | Yes | Yes | Py | No | Py | No | N/A | N/A | N/A | Yes | Yes | N/A | Yes | Low |
| *De Souto Barreto* | Yes | Yes | Yes | Py | Yes | Yes | Yes | Yes | Yes | N/A | No | Yes | N/A | N/A | Yes | Yes | Yes | Yes | High |
| *Pimouguet* | No | No | No | No | No | No | No | Py | No | N/A | No | N/A | N/A | N/A | No | No | N/A | No | Critically low |
| *Dixon* | Yes | No | No | Yes | Yes | No | Py | Py | No | No | No | N/A | N/A | N/A | Yes | No | N/A | Yes | Critically low |
| *Tam-Tham* | Yes | Yes | Yes | Py | Yes | Yes | Yes | Yes | Yes | Py | No | Yes | N/A | Yes | Yes | Yes | Yes | Yes | Moderate |
| *Frost* | Yes | No | Yes | Yes | Yes | Yes | Yes | Py | Py | Py | No | Yes | N/A | Yes | Yes | Yes | No | Yes | Critically low |
| *Phelan* | Yes | No | No | Py | Yes | Yes | Yes | Py | No | N/A | No | N/A | N/A | N/A | Yes | Yes | N/A | Yes | Critically low |
| *Quinn* | Yes | Yes | No | Py | Yes | Yes | Yes | Py | Yes | N/A | No | Yes | N/A | Yes | Yes | Yes | Yes | Yes | Moderate |
| *Lee* | Yes | No | Yes | Yes | Yes | Yes | Yes | Yes | Yes | N/A | No | Yes | N/A | Yes | Yes | Yes | Yes | Yes | Low |
| *Reilly* | Yes | Yes | Yes | Yes | Yes | Yes | Yes | Yes | Yes | N/A | Yes | Yes | N/A | Yes | Yes | Yes | Yes | Yes | High |
| *Ma* | Yes | No | Yes | Py | Yes | Yes | Py | Py | No | No | No | N/A | N/A | N/A | Yes | No | N/A | Yes | Critically low |
| ***Sawan*** | Yes | No | Yes | Py | Yes | Yes | Yes | Py | N/A | Yes | No | N/A | N/A | N/A | No | Yes | N/A | Yes | Critically low |
| ***Demanes*** | No | No | No | No | No | No | Py | Yes | Py | N/A | No | N/A | N/A | N/A | No | No | N/A | Yes | Critically low |
| ***McCausland*** | Yes | Py | Yes | Yes | Yes | Yes | Yes | Py | No | Py | No | N/A | N/A | N/A | Yes | Yes | N/A | Yes | Moderate |
| ***Hovsepian*** | Yes | No | Yes | Py | Yes | Yes | Yes | Py | Yes | Yes | No | N/A | N/A | N/A | Yes | Yes | N/A | Yes | High |
| ***Tunnard*** | Yes | Py | Yes | Py | Yes | Yes | Yes | Yes | Py | Py | No | N/A | N/A | N/A | Yes | Yes | N/A | Yes | High |
| ***Feast*** | Yes | Py | Yes | Yes | No | No | Yes | Yes | Yes | Py | No | N/A | N/A | N/A | Yes | Yes | N/A | Yes | High |
| ***Packer*** | Yes | No | Yes | Yes | Yes | Yes | Py | Yes | Yes | N/A | No | Yes | N/A | Yes | Yes | Yes | No | Yes | Critically low |
| ***Godard-Sebillotte*** | Yes | Py | No | Py | Yes | Yes | Yes | Yes | No | N/A | No | Yes | N/A | Yes | Yes | Yes | Yes | Yes | Low |
| ***Casarez*** | Yes | Py | Yes | Py | Yes | No | Yes | Py | Py | Py | No | N/A | N/A | N/A | Yes | No | N/A | Yes | Moderate |
| ***Khanassov*** | No | No | Yes | Yes | Yes | Yes | Yes | Yes | Py | Py | No | N/A | N/A | N/A | Yes | No | N/A | Yes | Low |
| ***Andrews*** | Yes | Yes | Yes | Py | Yes | Yes | Yes | Py | Yes | Yes | No | N/A | N/A | N/A | Yes | Yes | N/A | Yes | Moderate |
| ***Bocks*** | Yes | Py | Yes | Yes | Yes | Yes | Yes | Yes | No | No | No | N/A | N/A | N/A | No | Yes | N/A | Yes | Critically low |
| ***Lee (2025)*** | Yes | Yes | Yes | Py | Yes | Yes | Yes | Py | Yes | Py | No | N/A | N/A | N/A | Yes | Yes | N/A | Yes | Moderate |
| ***Smith*** | Yes | No | Yes | Py | Yes | Yes | Yes | Py | N/A | Yes | No | N/A | N/A | N/A | No | No | N/A | Yes | Critically low |

Py – partial yes; N/A – no meta-analysis performed
\*Critical domain

**Figure 1:**
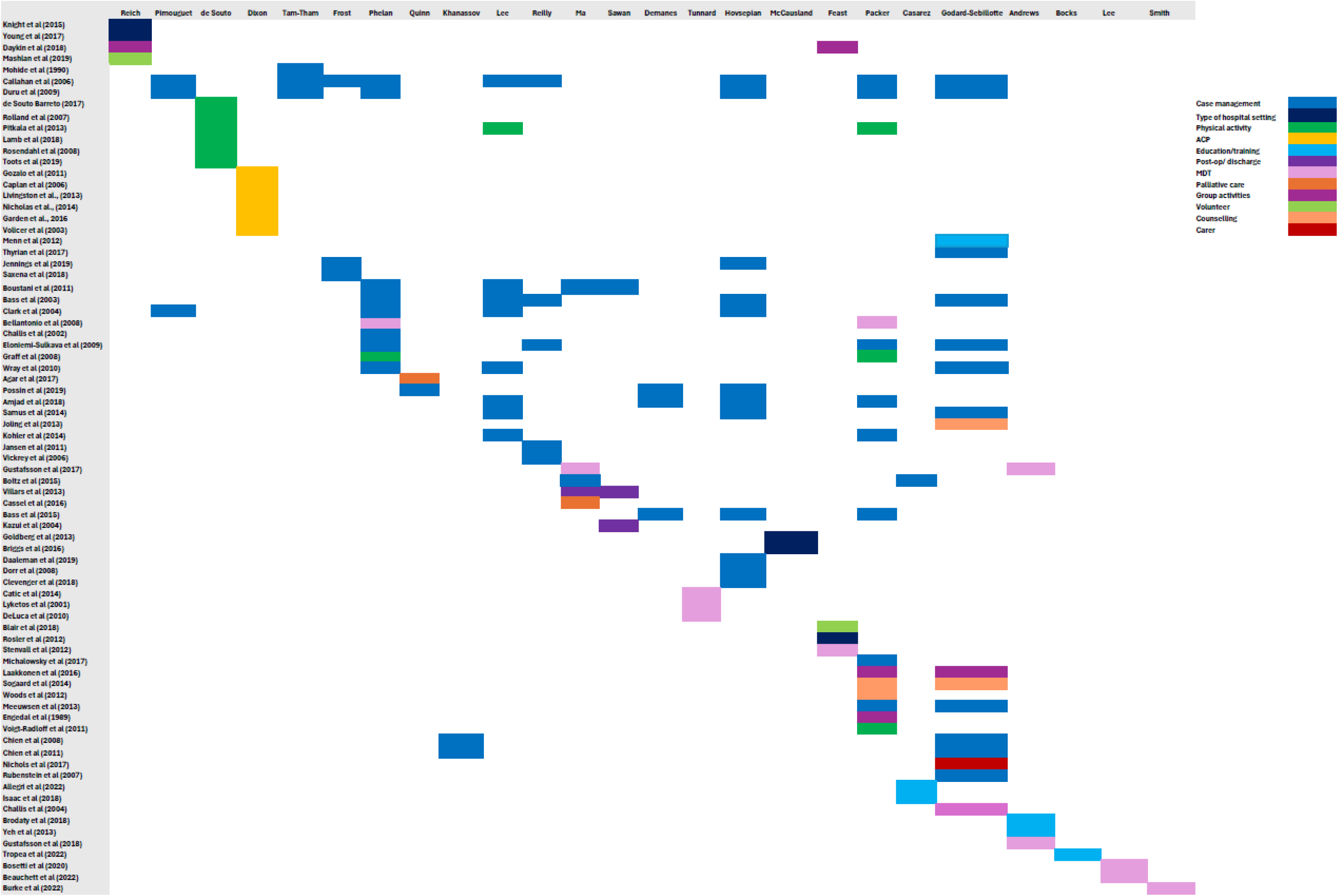
Citation matrix.

**Table 3:** PRIOR checklist.

| Section Topic | # | Item | Location reported |
| --- | --- | --- | --- |
| TITLE |  |  |  |
| Title | 1 | Identify the report as an overview of reviews. | Pg 1 |
| ABSTRACT |  |  |  |
| Abstract | 2 | Provide a comprehensive and accurate summary of the purpose, methods, and results of the overview of reviews. | Pg 2 |
| INTRODUCTION |  |  |  |
| Rationale | 3 | Describe the rationale for conducting the overview of reviews in the context of existing knowledge. | Pg 3 |
| Objectives | 4 | Provide an explicit statement of the objective(s) or question(s) addressed by the overview of reviews. | Pg 3 |
| METHODS |  |  |  |
| Eligibility criteria | 5a | Specify the inclusion and exclusion criteria for the overview of reviews. If supplemental primary studies were included, this should be stated, with a rationale. | Pg 4 |
|  | 5b | Specify the definition of 'systematic review' as used in the inclusion criteria for the overview of reviews. | Pg 4 |
| Information sources | 6 | Specify all databases, registers, websites, organizations, reference lists, and other sources searched or consulted to identify systematic reviews and supplemental primary studies (if included). Specify the date when each source was last searched or consulted. | Pg 3 |
| Search strategy | 7 | Present the full search strategies for all databases, registers and websites, such that they could be reproduced. Describe any search filters and limits applied. | Pg 3 |
| Selection process | 8a | Describe the methods used to decide whether a systematic review or supplemental primary study (if included) met the inclusion criteria of the overview of reviews. | Pg 4 |
|  | 8b | Describe how overlap in the populations, interventions, comparators, and/or outcomes of systematic reviews was identified and managed during study selection. | Pg 4 |
| Data collection process | 9a | Describe the methods used to collect data from reports. | Pg 4 |
|  | 9b | If applicable, describe the methods used to identify and manage primary study overlap at the level of the comparison and outcome during data collection. For each outcome, specify the method used to illustrate and/or quantify the degree of primary study overlap across systematic reviews. | Pg 4 |
|  | 9c | If applicable, specify the methods used to manage discrepant data across systematic reviews during data collection. | Pg 4 |
| Data items | 10 | List and define all variables and outcomes for which data were sought. Describe any assumptions made and/or measures taken to identify and clarify missing or unclear information. | Pg 4 |
| Risk of bias assessment | 11a | Describe the methods used to <u>assess</u> risk of bias or methodological quality of the included systematic reviews. | Pg 5 |
|  | 11b | Describe the methods used to <u>collect</u> data on (from the systematic reviews) and/or <u>assess</u> the risk of bias of the primary studies included in the systematic reviews. Provide a justification for instances where flawed, incomplete, or missing assessments are identified but not re-assessed. | Pg 5 |
|  | 11c | Describe the methods used to <u>assess</u> the risk of bias of supplemental primary studies (if included). |  |
| Synthesis methods | 12a | Describe the methods used to summarize or synthesize results and provide a rationale for the choice(s). | Pg 5 |
|  | 12b | Describe any methods used to explore possible causes of heterogeneity among results. |  |
|  | 12c | Describe any sensitivity analyses conducted to assess the robustness of the synthesized results. |  |
| Reporting bias assessment | 13 | Describe the methods used to <u>collect</u> data on (from the systematic reviews) and/or <u>assess</u> the risk of bias due to missing results in a summary or synthesis (arising from reporting biases at the levels of the systematic reviews, primary studies, and supplemental primary studies, if included). | Pg 5 |
| Certainty assessment | 14 | Describe the methods used to <u>collect</u> data on (from the systematic reviews) and/or <u>assess</u> certainty (or confidence) in the body of evidence for an outcome. | Pg 5 |
| RESULTS |  |  |  |
| Systematic review and supplemental primary study selection | 15a | Describe the results of the search and selection process, including the number of records screened, assessed for eligibility, and included in the overview of reviews, ideally with a flow diagram. | Pg 5 |
|  | 15b | Provide a list of studies that might appear to meet the inclusion criteria, but were excluded, with the main reason for exclusion. | Pg 6 |
| Characteristics of systematic reviews and supplemental primary studies | 16 | Cite each included systematic review and supplemental primary study (if included) and present its characteristics. | Pg 6 |
| Primary study overlap | 17 | Describe the extent of primary study overlap across the included systematic reviews. | Pg 6 |
| Risk of bias in systematic reviews, primary studies, and supplemental primary studies | 18a | Present assessments of risk of bias or methodological quality for each included systematic review. | Pg 14 |
|  | 18b | Present assessments (<u>collected</u> from systematic reviews or <u>assessed</u> anew) of the risk of bias of the primary studies included in the systematic reviews. | Pg 14 |
|  | 18c | Present assessments of the risk of bias of supplemental primary studies (if included). |  |
| Summary or synthesis of results | 19a | For all outcomes, summarize the evidence from the systematic reviews and supplemental primary studies (if included). If meta-analyses were done, present for each the summary estimate and its precision and measures of statistical heterogeneity. If comparing groups, describe the direction of the effect. | Pg 14-26 |
|  | 19b | If meta-analyses were done, present results of all investigations of possible causes of heterogeneity. |  |
|  | 19c | If meta-analyses were done, present results of all sensitivity analyses conducted to assess the robustness of synthesized results. |  |
| Reporting biases | 20 | Present assessments (*collected* from systematic reviews and/or *assessed* anew) of the risk of bias due to missing primary studies, analyses, or results in a summary or synthesis (arising from reporting biases at the levels of the systematic reviews, primary studies, and supplemental primary studies, if included) for each summary or synthesis assessed. | Pg 14 |
| Certainty of evidence | 21 | Present assessments (*collected* or *assessed* anew) of certainty (or confidence) in the body of evidence for each outcome. | Pg 14 |
| DISCUSSION |  |  |  |
| Discussion | 22a | Summarize the main findings, including any discrepancies in findings across the included systematic reviews and supplemental primary studies (if included). | Pg 27 |
|  | 22b | Provide a general interpretation of the results in the context of other evidence. | Pg 27/28 |
|  | 22c | Discuss any limitations of the evidence from systematic reviews, their primary studies, and supplemental primary studies (if included) included in the overview of reviews. Discuss any limitations of the overview of reviews methods used. | Pg 27 |
|  | 22d | Discuss implications for practice, policy, and future research (both systematic reviews and primary research). Consider the relevance of the findings to the end users of the overview of reviews, e.g., healthcare providers, policymakers, patients, among others. | Pg 28 |
| OTHER INFORMATION |  |  |  |
| Registration and protocol | 23a | Provide registration information for the overview of reviews, including register name and registration number, or state that the overview of reviews was not registered. | Pg 2/3 |
|  | 23b | Indicate where the overview of reviews protocol can be accessed, or state that a protocol was not prepared. | Pg 2/3 |
|  | 23c | Describe and explain any amendments to information provided at registration or in the protocol. Indicate the stage of the overview of reviews at which amendments were made. | Pg 3 |
| Support | 24 | Describe sources of financial or non-financial support for the overview of reviews, and the role of the funders or sponsors in the overview of reviews. | Pg 30 |
| Competing interests | 25 | Declare any competing interests of the overview of reviews' authors. | Pg 30 |
| Author information | 26a | Provide contact information for the corresponding author. | Pg 1 |
|  | 26b | Describe the contributions of individual authors and identify the guarantor of the overview of reviews. | Pg 30/21 |
| Availability of data and other materials | 27 | Report which of the following are available, where they can be found, and under which conditions they may be accessed: template data collection forms; data collected from included systematic reviews and supplemental primary studies; analytic code; any other materials used in the overview of reviews. | Pg 30 |

